# A cross-sectional study of patients presenting for hospital-based screening for COVID-19: risk of disease, and healthcare access preferences

**DOI:** 10.1101/2020.04.15.20067207

**Authors:** Amanda Rojek, Martin Dutch, Daniel Peyton, Rachel Pelly, Mark Putland, Harriet Hiscock, Jonathan Knott, on behalf of the RMH-ED COVID-19 Working Group

## Abstract

**Introduction:** Early during the SARS-CoV-2 pandemic, Australian emergency departments (EDs) have experienced an unprecedented surge in patients seeking screening for COVID-19. Understanding what proportion of these patients require screening, who can be safely screened in community based models of care, and who requires an emergency departments to care for them is critical for workforce and infrastructure planning across the healthcare system, as well as public messaging campaigns.

**Methodss:** In this cross sectional survey, we screened patients presenting to a SARS-CoV-2 screening clinic in a tertiary hospital Emergency Department in Melbourne, Australia. We assessed the proportion of patients who met screening criteria; self-reported symptom severity; reasons why they came to the ED for screening; views on community-based models of care; and sources of information accessed about COVID-19.

**Results:** We included findings from 1846 patients who presented to the Emergency Department (ED) for COVID-19 screening from 18th to 30th March 2020. Most patients (55.3%) did not meet criteria for screening and most (57.6%) had mild or no (13.4%) symptoms. The main reason for coming to the ED was being referred by a telephone health service (31.3%) and 136 (7.4%) said they tried to contact their GP but could not get an appointment. Only 47 (2.6%) said they thought the disease was too specialized for their GP to manage. Patients accessed numerous information sources, commonly government websites (68.4%) and other websites (51.3%) for COVID-19 information.

**Conclusions:** if we are to ensure that emergency departments can cope with the likely surge in presentations requiring resuscitation or inpatient care COVID-19, we should strengthen access to alternative services to triage patients to prevent unnecessary presentations at health services, and to direct those who are well but require screening away from EDs.

## 1 Introduction

The Coronavirus Disease 2019 (COVID-19) pandemic, caused by Severe Acute Respiratory Syndrome Coronavirus 2 (SARS-CoV-2) risks placing “*overwhelming demands on our health system*”.(1-4) This is particularly so for hospital Emergency Departments (EDs) and General Practitioners (GPs) who are seen as key providers of screening for the virus.(5)

However, during the early phase of importation of SARS-CoV-2 in Australia there was limited community transmission.(6) Under this scenario, the pre-test probability of infection in individuals without epidemiological risk (such as relevant travel, or close contact with a confirmed case) is low. Unnecessary presentations for testing during this phase places additional strain on the public health service, including EDs. As we enter the pandemic phase of SARS-CoV-2, it will become critically important that the right patient is assessed, and then if necessary, treated at the right facility. This can be achieved by considering alternative strategies for screening well patients, and managing mild cases.(5)

There is a paucity of literature describing the healthcare-seeking behaviours of those who present for screening for a high consequence infectious disease in Australia. As Australia moves from containment to mitigation of COVID-19, we must rapidly generate evidence to calibrate and direct policy for safe, effective and efficient models of care for COVID-19. We do not know how many mildly unwell patients elect to present to an ED for screening and the barriers and facilitators to other models of care.

Here, we provide the first characterisation of patients presenting to hospital-based screening early during the outbreak.

## 2 Methods

### Setting

The Royal Melbourne Hospital (RMH) is a metropolitan tertiary hospital service, with an emergency department that manages 80 000 patients per year. Co-located with this emergency department, a SARS-CoV-2 screening clinic was established on 25/01/2020 to deal with the surge in patients presenting for screening. (7)

### Design and Participants

A consecutive sample of patients presenting to the SARS-CoV-2 screening clinic from 18/03/2020 to 30/03/2020 were invited to complete a brief survey as part of their clinic registration process. Patients were included if they were aged 16 years or older, able to complete the self-registration survey independently, or communicate to a staff member who could complete it on their behalf. Patients were excluded if they required urgent medical treatment (as assessed by triage staff) because they were re-streamed from the screening are into the resuscitation space of the ED.

Patients completed the study-designed survey embedded into a larger epidemiological and clinical triage system (described elsewhere) (7) upon registering at the clinic. The web-based (REDCap™) survey was completed either on their phone or on a hospital supplied tablet. When required, patients were assisted by triage staff to help them complete the survey, including assisting with language translation. Survey questions are outlined in Table 2.

**Table 1.**
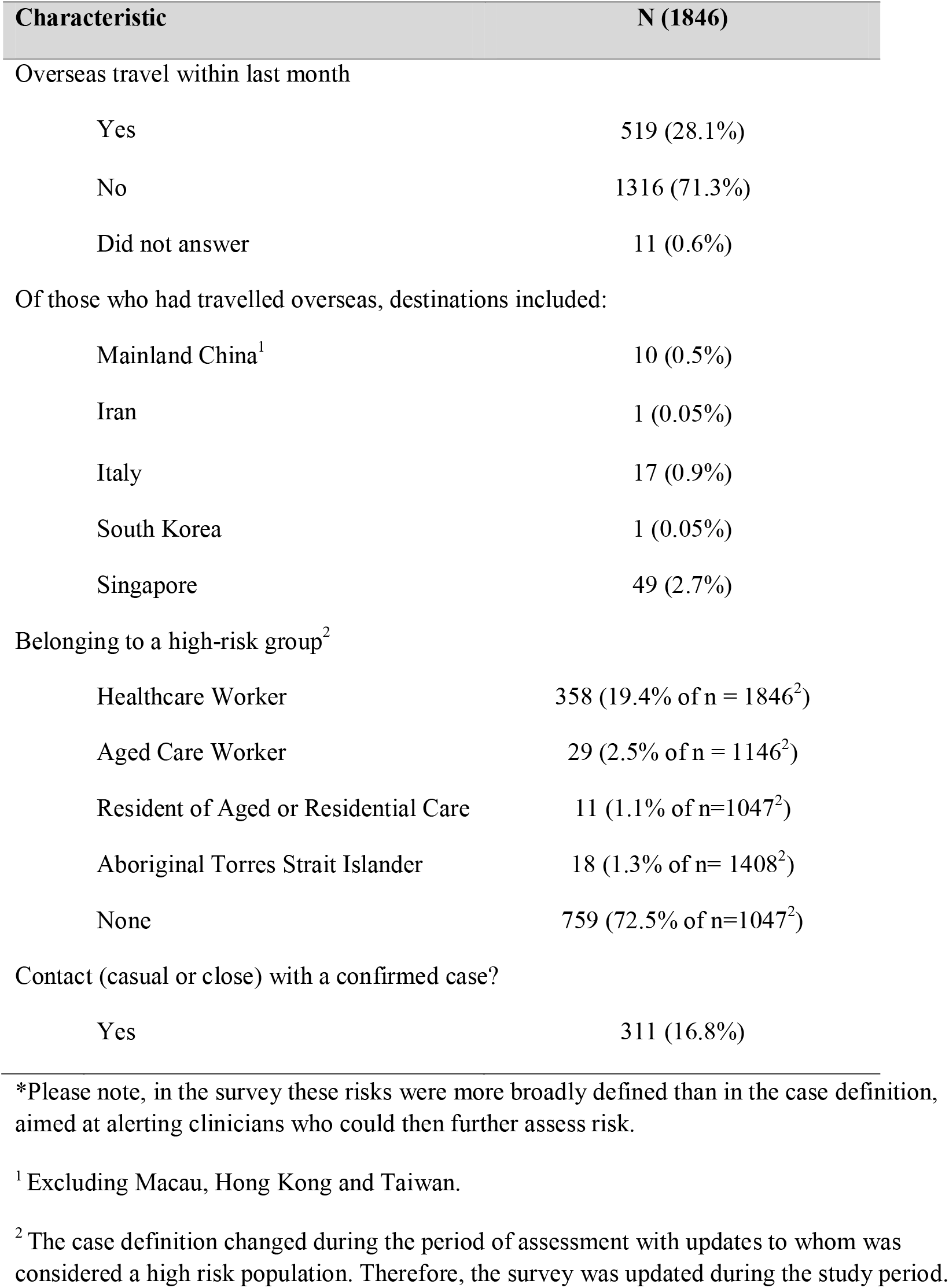
Epidemiological risk factors* of survey population

| Characteristic | N (1846) |
| --- | --- |
| Overseas travel within last month |  |
| Yes | 519 (28.1%) |
| No | 1316 (71.3%) |
| Did not answer | 11 (0.6%) |
| Of those who had travelled overseas, destinations included: |  |
| Mainland China<sup>1</sup> | 10 (0.5%) |
| Iran | 1 (0.05%) |
| Italy | 17 (0.9%) |
| South Korea | 1 (0.05%) |
| Singapore | 49 (2.7%) |
| Belonging to a high-risk group<sup>2</sup> |  |
| Healthcare Worker | 358 (19.4% of n = 1846<sup>2</sup>) |
| Aged Care Worker | 29 (2.5% of n = 1146<sup>2</sup>) |
| Resident of Aged or Residential Care | 11 (1.1% of n=1047<sup>2</sup>) |
| Aboriginal Torres Strait Islander | 18 (1.3% of n= 1408<sup>2</sup>) |
| None | 759 (72.5% of n=1047<sup>2</sup>) |
| Contact (casual or close) with a confirmed case? |  |
| Yes | 311 (16.8%) |

\*Please note, in the survey these risks were more broadly defined than in the case definition, aimed at alerting clinicians who could then further assess risk.
<sup>1</sup> Excluding Macau, Hong Kong and Taiwan.
<sup>2</sup> The case definition changed during the period of assessment with updates to whom was considered a high risk population. Therefore, the survey was updated during the study period.

**Table 2.**
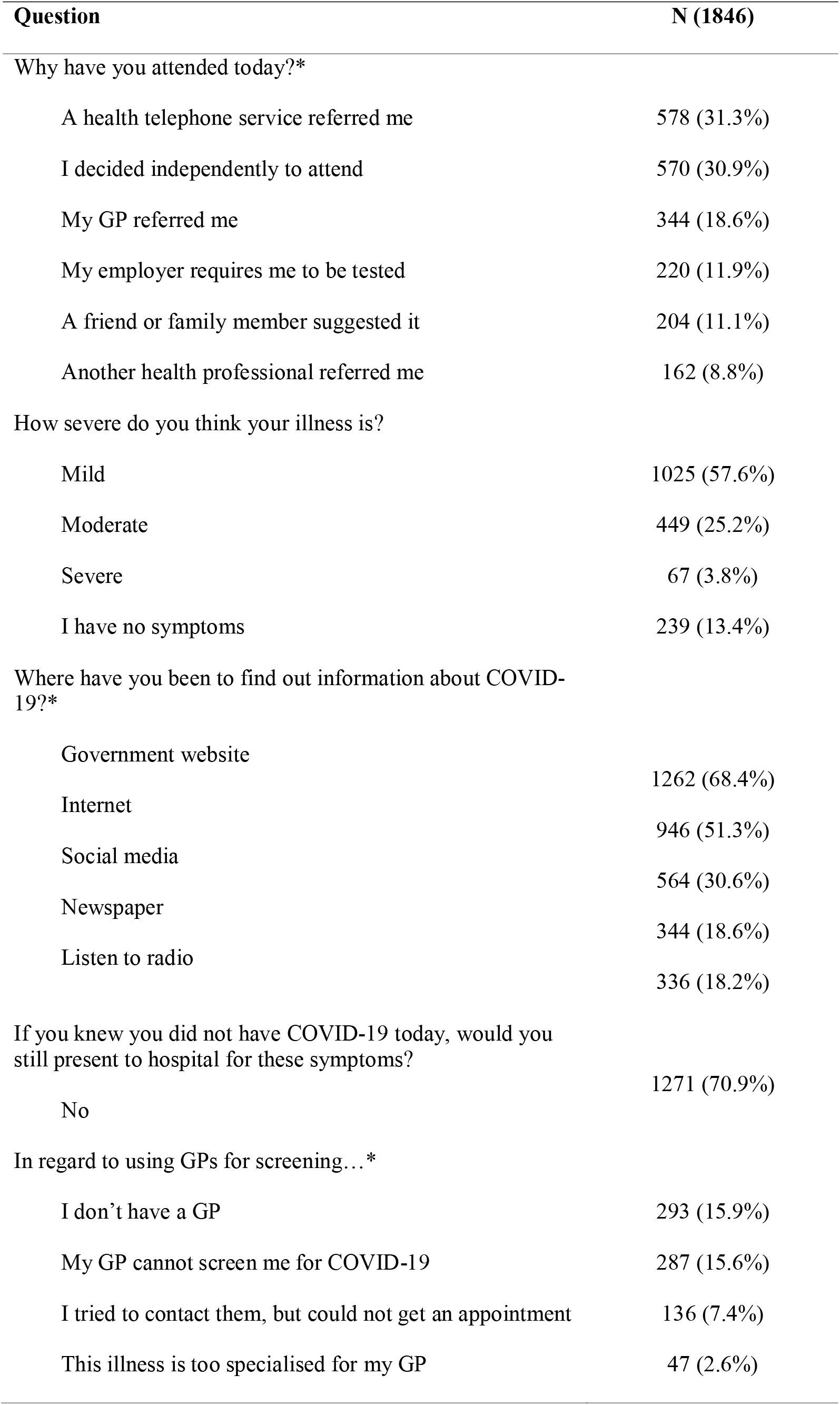
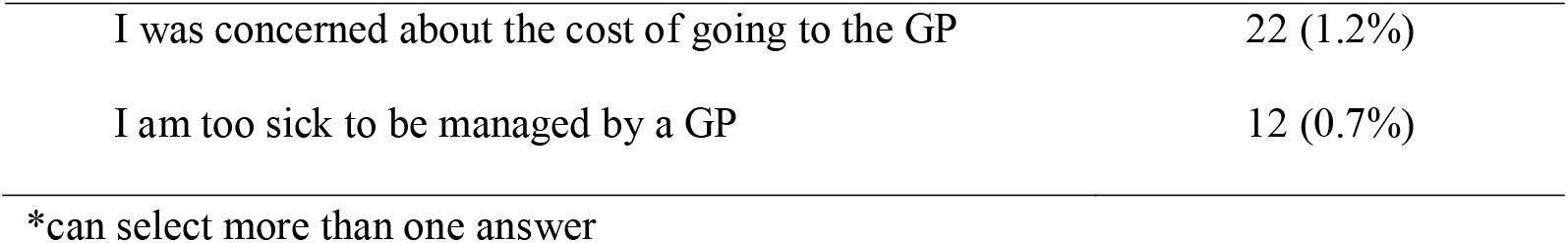
Health access preference questions for patients presenting for screening

Data were stored on REDCap via the hospital’s server in accordance with relevant information security protocols.. This quality assurance project (QA202025 RMH Ethics) was approved by Melbourne Health (05/03/2020).

### Data Cleaning and Analysis

Data was cleaned and analysed using Stata (version 15.0; College Station, Texas, USA). Survey responses completed for children (i.e., age under 16-years) were excluded from analyses (*n* = 3). Date of birth and age was excluded from analyses if the response was impossible (e.g., age of 1000-years). It was assumed that the date of birth was simply input incorrectly at the time of data collection and, accordingly, all other data from those surveys was included in the analysis.

Descriptive data are summarised with median and inter-quartile range for skewed data whilst categorical data are described with overall frequency and proportions. Where analysis refers to the suspected case definition, this refers to the prevailing Victorian Department of Health and Human Services (DHHS) case definition for COVID-19.(8)

## 3 Results

2359 patients arrived at the RMH and requested screening for COVID of whom 1846 (78.3%) completed the survey. Most were female (51.3%) with a median age of 32 years (interquartile range 25.5-42.9 years).

### Risk of COVID-19

Whether a patient required screening depended on their travel history, contact with a confirmed case, or belonging to a high-risk group (variably defined due to changing case definitions) (see Table 1). Of our cohort, 825 of the 1846 (44.7%) patients met criteria for screening. The proportion of patients each day who met the criteria for COVID-19 testing did not differ greatly or show any discernible pattern over time (see Figure 1).

**Figure 1.**
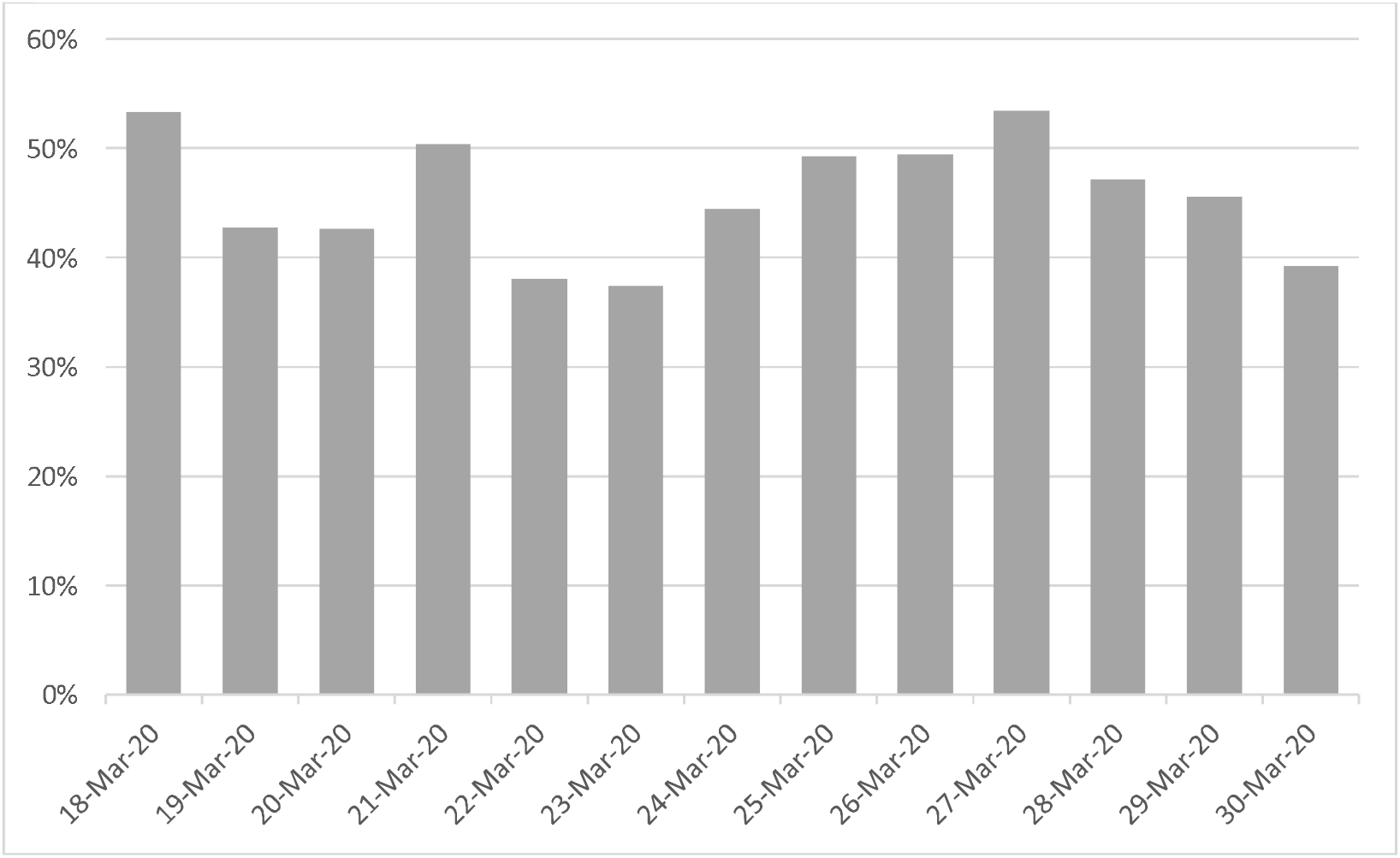
Proportion of patients over time who met criteria for COVID-19 testing.

### Self-assessed severity of illness

Most patients (1025; 57.6%) rated their illness as only mild, with another 239 (13.4%) stating they had no symptoms at all (see Table 2). A minority of presenting patients self-reported moderate (449; 25.2%) or severe (67; 3.8%) symptoms. The majority (1271; 70.9%) of patients would not have presented to hospital for their symptoms if they knew they did not have COVID-19.

### Healthcare access preferences

When asked about utilising other models of care for screening for COVID-19, some patients (287; 15.6%) reported that their GP could not screen them for COVID-19 and 293 (15.9%) patients who stated they did not have a GP (see Table 2). A minority of patients did not use their GP for screening because of the cost (22; 1.2%) or because they thought that the illness was too specialised for care by a GP (47; 2.6%).

For these patients presenting to ED for COVID-19 screening, the survey explored the catalyst for them choosing to attend (See Table 2). Most commonly, either a telephone health service referred them (578; 31.3%) or patients presented after making an independent decision to do so (570; 30.9%). Other reasons for attending included being referred by a GP (344; 18.6%) or an employer required the patient to be tested (220; 11.9%).

Patients had accessed a variety of sources for information about COVID-19, primarily accessing information from government websites (1262; 68.4%), the internet (946; 51.3%) and social media (564; 30.6%).

## 4 Discussion

This data provides the first contemporary review of the patients presenting to a tertiary ED for COVID-19 screening during a period before widespread community transmission. Of note, most patients had either mild or no symptoms and would not have presented to the ED had they known they did not have COVID-19. Of those who did present, a significant proportion (55.3%) did not meet screening criteria. Further, few patients presenting to the screening clinic thought that their illness was either too severe, or too specialised, for their GP to manage. Patients had sought information from a variety of sources about COVID-19.

To our knowledge, this is the first study to present the clinical characteristics and help-seeking behaviours of patients attending an Australian ED for COVID-19 screening. We had a good (78%) response rate and included non-English speaking patients. However, our study has some limitations. The survey was conducted in a single ED with only those who had presented to the ED (and not other healthcare providers such as GPs) so our results may not generalise to patient groups beyond our setting. Further, we excluded patients who were assessed at triage to require active medical treatment or resuscitation, which may result in an under-representation of the true number of severe cases presenting to the ED for COVID-19 screening. The survey was based on self-report and we were unable to verify patient responses (e.g. if they had tried to but been unable to get an appointment with a GP).

Our data have several clinical and policy implications. Ideally, only patients who require COVID-19 testing based on epidemiological or clinical history should present for screening, with perhaps only those who are unwell presenting to EDs. Given that most patients who presented to our ED had mild symptoms or did not meet screening criteria, serious consideration must be given to re-directing such patients to an alternate service. Failing to do so *before* patients present to an ED will almost certainly result in EDs becoming overwhelmed, given the anticipated upcoming surge of unwell patients presenting with COVID-19 related respiratory failure.

Re-directing well patients or those with only mild illness will require accurate triaging as well as alternative services. Possibilities for alternate services to conduct triaging incorporating epidemiological and clinical histories include a hotline, community-based screening service, GP practices, or self-screening via an app/online service. Those patients who do warrant testing for SARS-CoV-2 could then be directed to an appropriate service, with well patients being directed to a community clinic or general practice and unwell patients being referred to ED with their specialised services in critical care. Our data suggest patients are willing to be screened in the community, as demonstrated by the small proportion of patients who reported that they thought the illness was too specialised for GPs to manage.

Although we collected data over a brief period of time, we saw no decline in the proportion of patients presenting who did not meet criteria for testing, despite widespread public messaging about screening criteria at the time. Given that patients in this study had accessed a variety of sources for COVID-19 information, consistent triaging and messaging across these services about who should/should not be screened will be vital to minimise unnecessary screening.

## 5 Conclusion

The COVID-19 pandemic is likely to place an unprecedented and sustained surge of patients to emergency departments around Australia. In order to avoid additional burden on the public health system, it is imperative the right patient is treated at the right facility. During this pandemic phase, there is an urgent need to explore alternative avenues to triage and if needed, screen these predominantly well patients, such as through community clinics or general practices. This will ensure the right patient is treated at the right facility.

## Data Availability

The data that support the findings of this study are available from the corresponding author, upon reasonable request.

## 7 Acknowledgements

RMH ED COVID-19 Working Group: Nicola Walsham, Ben Smith, David Camilleri, Susan Harding, Steve Pincus, Emma Gardiner, Emma Smith, Naomi Watson, Bruce Garbutt, Cherylnn McGurgan, Elizabeth Bradbury. The Murdoch Children’s Research Institute is funded through Victorian Operating Fund. HH holds a NHMRC Practitioner fellowship grant GN1136222.

